# Maternal MTHFR A1298C polymorphism and risk of congenital heart disease in fetus

**DOI:** 10.1101/19010298

**Authors:** Vandana Rai

## Abstract

Methylenetetrahydrofolate reductase (MTHFR) is an important enzyme involved in folate metabolism, DNA synthesis and methylation. A number of studies have examined the association of maternal MTHFR A1298C polymorphism with congenital heart disease (CHD) susceptibility; however, the conclusions were contradictory. To clarify the influence of maternal MTHFR A1298C polymorphism on CHD, a meta-analysis of seventeen case-control studies was carried out. Four electronic databases - Pubmed, Google Scholars, Elsevier and Springer Link were searched upto June, 2018 for suitable articles. The pooled odds ratios (ORs) with 95% confidence intervals (95% CIs) were used to evaluate the association. Meta-analysis was performed by Mix and MetaAnalyst programs. The results of meta-analysis suggested that except co-dominant model, maternal A1298C polymorphism is risk for CHD in fetus using overall comparisons in four genetic models (C vs. A: OR= 1.19, 95% CI= 1.00-1.41, p= 0.04; CC+AC vs. AA: OR= 1.19, 95% CI= 0.97-1.4, p= 0.04; CC vs. AA: OR= 1.46, 95% CI= 1.00-2.13, p= 0.04; AC vs. AA OR= 1.13, 95% CI=0.93-1.36, p= 0.23; CC vs. AC+AA: OR=1.34, 95% CI=1.1-1.6, p=0.01). Publication bias was absent using four genetic models. In conclusion, results of present meta-analysis showed significant association between maternal MTHFR A1298C polymorphism and CHD risk.

## Introduction

Cardiac malformations rank among the most common congenital malformations. The incidence of congenital heart disease (CHD) is about 8 per 1000 live birth (Mitchell et al., 1971; Hofmann and Christianson, 1978; Winter et al.,2007) and are mainly due to incomplete development of the heart during pregnancy (Bozovic et al., 2011). Their origin is considered to be multifactorial, resulting from an interaction between genetic predisposition and environmental factors. Periconceptional folic acid, reduces the risk of CHD like -neural tube defects (NTD) (Czeizel and Dudas, 1992). Although the mechanism by which folic acid exerts its protective effect is unclear, the teratogenic process resulting from folate insufficiency may be related to hyperhomocysteinemia (Nelen, 2001; Vollset et al., 2000). Methylenetetrahydrofolate reductase (MTHFR) gene polymorphism is an important genetic factor for hyperhomocysteinemia (Sibani et al.,2000).

MTHFR is a critical enzyme in folate and methionine metabolism. It irreversibly catalyzes the reduction of 5,10- methylene-tetrahydrofolate to 5-methyltetrahydrofolate, which one is the methyl donor for the conversion of homocysteine to methionine (Goyette et al., 1994). Several folate pathway genes showed single nucleotide polymorphism (SNPs) and frequency of these polymorphisms varies greatly word wide (Rai et al.,2010,2012a,b,2013; Yadav et al., 2017). The MTHFR gene is located on chromosome 1p36.3. Among all the identified polymorphisms, C677T and A1298C polymorphisms are clinically important and variant MTHFR enzymes (Ala222Val and Glu429ALa) are associated with reduced enzyme activity and hyperhomocysteinemia (Frosst et al.,1995; Weisberg et al.,1998; Rozen,1997).

A1298C polymorphism involves A to C nucleotide substitution at 1298th position (Weisberg et al. 1998), leading to a glutamate to alanine substitution (Glu429Ala) in the MTHFR protein. Glutamate to alanine amino acid substitution at 429th position in MTHFR enzyme, reduces 40% enzyme activity. A1298C allele frequency differs greatly in various ethnic groups of the world. The prevalence of the A1298C homozygote variant genotype ranges from 7 to 12% in White populations from North America and Europe and lower frequencies have been reported in Hispanics (4 to 5 %) and Asian populations (1 to 4%) (Botto and Young, 2000). MTHFR A1298C gene polymorphism is associated with a number of diseases like- Down syndrome (Rai et al., 2006), neural tube defects (van der Put et al., 1997), cleft lip and palate (Chorna et al.,20011) and psychiatric disorders (Sazci et al., 2005) etc. Numerous case control studies were investigated the association between maternal MTHFR A1298C polymorphism and CHD but the results were contradictory. Hence, we performed meta-analysis of case control studies to conclude the role of maternal MTHFR A1298C polymorphism in fetal CHD risk.

## Methods

Article search was carried out in four electronic databases (Pubmed, Google scholar, Elsevier and Springer Link) up to June, 2018 using key terms - ‘methylenetetrahydrofolate reductase or MTHFR’, ‘A1298C’, ‘polymorphism’ and ‘congenital heart disease or CHD’. Reference list of eligible articles were also searched for relevant studies.

### Data selection

Following data from each included article were extracted: author name; ethnicity; country of origin; journal name; year of publication; number of cases and controls; and number of MTHFR A1298C genotypes in mothers of CHD cases and mothers of healthy children as controls. **Inclusion and exclusion criteria:**

A strict inclusion and exclusion criteria was followed to select the studies for meta-analysis. Criteria for inclusion were as follows; (i) article should evaluated the association of maternal MTHFR A1298C gene polymorphism with CHD; (ii) article should be case-control association study; and (iii) the articles must report the sample size, distribution of alleles or genotypes for estimating the odds ratio (ORs) with 95% confidence interval (CIs). Studies were excluded if one of the following existed: (i) case-only studies, (ii) studies that contained duplicate data, and (iii) editorial, case reports or reviews.

### Statistical analysis

The present meta-analysis examined the overall association of maternal mutant C allele with the risk of CHD. The associations were indicated as odds ratios (ORs) with the corresponding 95% CI. The OR was estimated either by using fixed effects (Mantel and Haenszel, 1959) or random effects (DerSimonian and Laird, 1986) model depending upon heterogeneity. The heterogeneity between studies was tested using the Q-statistic and quantified by I^2^ statistic (Higgins et al., 2002).

### Sensitivity analysis

The distribution of the genotypes in the control group was tested for Hardy Weinberg equilibrium (HWE) and sensitivity analysis performed by exclusion of the studies in which control population was not in HWE.

### Publication bias

Publication bias was investigated by using funnel plots of standard error and precision. Egger’s regression intercept (Egger et al.,1997) was used to assess the publication bias. All p value were two sided, and p values <0.05 were considered statistically significant. All statistical analyses were performed using the computer program MIX version 1.7 (Bax et al., 2006) and MetaAnalyst (Wallace et al.,2013).

## Results

### Characteristics of included studies

Total seventeen studies (Storti et al.,2003; Nurk et al.,2004; Galdieri et al., 2007; Van Driel et al.,2008; Xu et al., 2010; Bozovic et al.,2011; Obermann-Borst et al.,2011; Weiner et al., 2012; Christense et al., 2013; Wang et al.,2013; Zidan et al.,2013; Huang et al.,2014; Sahiner et al.,2014; Li et al.,2015; Sayin et al.2015; Shi et al.,2015; Feng et al.,2016) were found suitable for the inclusion in present meta-analysis. The studies were published between 2003 and 2015. All these nine studies were performed in different countries- Brazil (Galdieri et al., 2007), China (Xu et al., 2010; Wang et al.,2013; Huang et al.,2014; Li et al.,2015; Sayin et al.2015; Shi et al.,2015; Feng et al.,2016), Croatia (Bozovic et al.,2011), Egypt (Zidan et al.,2013), Italy (Storti et al.,2003), Netherland (Obermann-Borst et al.,2011), Norway (Nurk et al.,2004; Van Driel et al.,2008), Russia (Weiner et al., 2011), Turkey (Sahiner et al.,2014), and USA (Christense et al., 2013). The lowest sample size was 25 (Nurk et al., 2004) and highest sample size was 502 (Xu et al., 20101) in included studies. Total number of cases were 2514 with genotype percentage of AA, AC and CC was 56.96%, 34.88% and 8.15% respectively. Total number of controls were 17,226 with genotypes percentage of AA, AC and CC were 47.53%, 42.51% and 9.95% respectively.

### Meta-analysis

In allele contrast meta-analysis, maternal mutant C allele showed significant association with CHD in fixed effect (OR= 1.14, 95%CI= 1.03-1.24, p= 0.007) and random effect (OR= 1.19, 95%CI= 0.95-1.36, p= 0.04) models (Table 2, Figure 1). Similar to allele contrast meta-analysis, pooled odds ratio for maternal mutant homozygote genotype (CC vs. AA) also showed significant association with CHD adopting both fixed (OR= 1.40, 95% CI= 1.11-1.46, p= 0.004) and random (OR= 1.46, 95%CI= 1.00-2.13, p= 0.04) effect models (Figure 2). Association of mutant heterozygous genotype (AC vs. AA; co-dominant model) did not show any association with CHD using fixed and random effect models. Dominant maternal mutant genotypes (CC+AC vs. AA; dominant model) showed significant association with CHD using both fixed (OR= 1.13; 95%CI= 1.0-1.26; p=0.04) and random (OR= 1.19; 95%CI= 0.97-1.4; p=0.04) effect models (Figure 3; Table 2). Meta-analysis using recessive genetic model (CC vs. AC+AA; recessive model) showed significant association with CHD with fixed (OR= 1.34; 95%CI= 1.1-1.6; p=0.01) and random (OR= 1.34; 95%CI= 0.99-1.81; p=0.28) effect models (Figure 4; Table 2).

**Table 1.** Distribution of different MTHFR A1298C genotypes in seventeen included meta-analysis

| Study | Population | Control | Case | Case genotype |  |  | Control Genotype |  |  | HWE P-value |
| --- | --- | --- | --- | --- | --- | --- | --- | --- | --- | --- |
|  |  |  |  | AA | AC | CC | AA | AC | CC |  |
| Storti et al.,2003 | Italy | 200 | 103 | 49 | 46 | 8 | 101 | 86 | 13 | 0.34 |
| Nurk et al.,2004 | Norway | 14474 | 25 | 9 | 13 | 3 | 6607 | 6342 | 1525 | 0.95 |
| Galdieri et al., 2007 | Brazil | 26 | 47 | 26 | 17 | 4 | 15 | 10 | 1 | 0.67 |
| Van Driel et al.,2008 | Norway | 251 | 230 | 104 | 102 | 24 | 116 | 104 | 31 | 0.31 |
| Xu et al.,2010 | China | 527 | 502 | 316 | 168 | 18 | 326 | 185 | 16 | 0.09 |
| Bozovic et al.,2011 | Croatia | 55 | 52 | 21 | 27 | 4 | 27 | 27 | 1 | 0.05 |
| Obermann-Borst et al.,2011 | Netherland | 183 | 139 | 69 | 57 | 13 | 75 | 90 | 18 | 0.23 |
| Weiner et al., 2012 | Russia | 362 | 48 | 33 | 13 | 2 | 168 | 152 | 42 | 0.39 |
| Christense et al., 2013 | USA | 65 | 182 | 98 | 71 | 13 | 36 | 22 | 7 | 0.21 |
| Zidan et al.,2013 | Egypt | 80 | 80 | 13 | 32 | 35 | 33 | 25 | 22 | 0.001 |
| Wang et al.,2013 | China | 188 | 170 | 115 | 45 | 10 | 133 | 47 | 8 | 0.22 |
| Huang et al.,2014 | China | 208 | 170 | 111 | 56 | 3 | 146 | 56 | 6 | 0.82 |
| Sahiner et al.,2014 | Turkey | 93 | 137 | 45 | 68 | 24 | 31 | 54 | 8 | 0.22 |
| Li et al.,2015 | China | 150 | 150 | 114 | 36 | 0 | 131 | 19 | 0 | 0.41 |
| Sayin et al.2015 | Turkey | 99 | 69 | 20 | 36 | 13 | 51 | 37 | 11 | 0.29 |
| Shi et al.,2015 | China | 216 | 153 | 95 | 39 | 19 | 157 | 53 | 6 | 0.055 |
| Feng et al.,2016 | China | 49 | 257 | 194 | 51 | 12 | 35 | 14 | 0 | 0.24 |

**Table 2:**
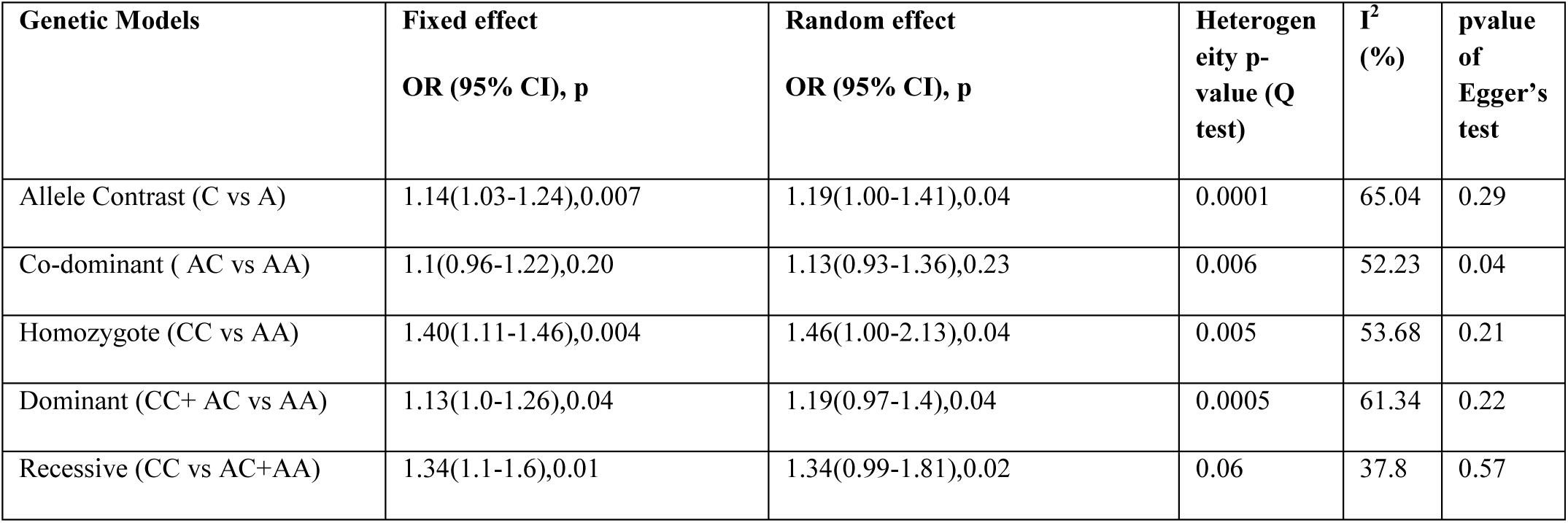
Summary estimates for the odds ratio (OR) of MTHFR A1298C in various allele/genotype contrasts, the significance level (p value) of heterogeneity test (Q test), and the I^2^ metric and publication bias p-value (Egger Test).

**Figure 1:**
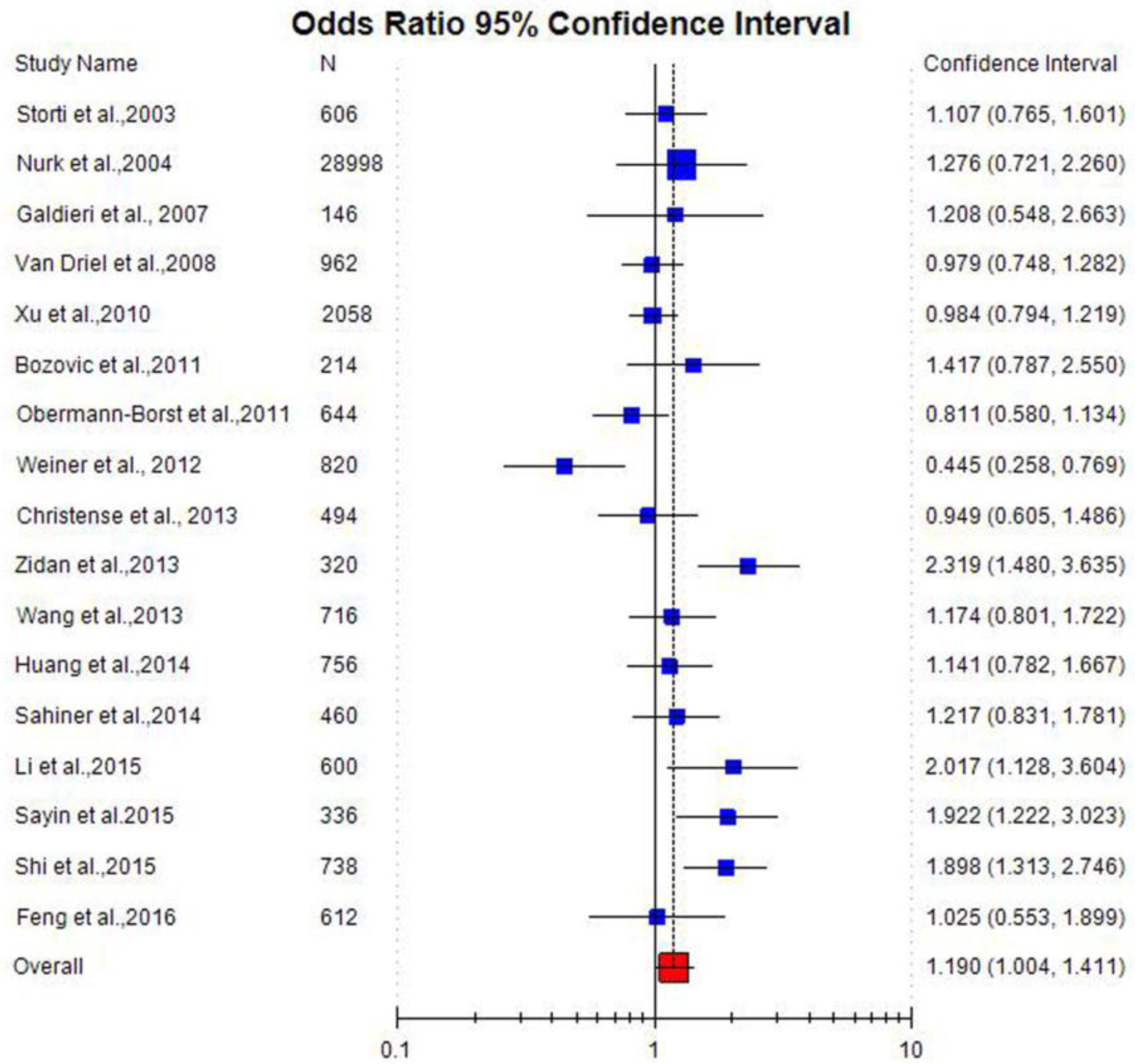
Forest plots (random effect) of allele contrast model (C versus A) showed significant association between maternal MTHFR A1298C polymorphism and CHD risk.

**Figure 2:**
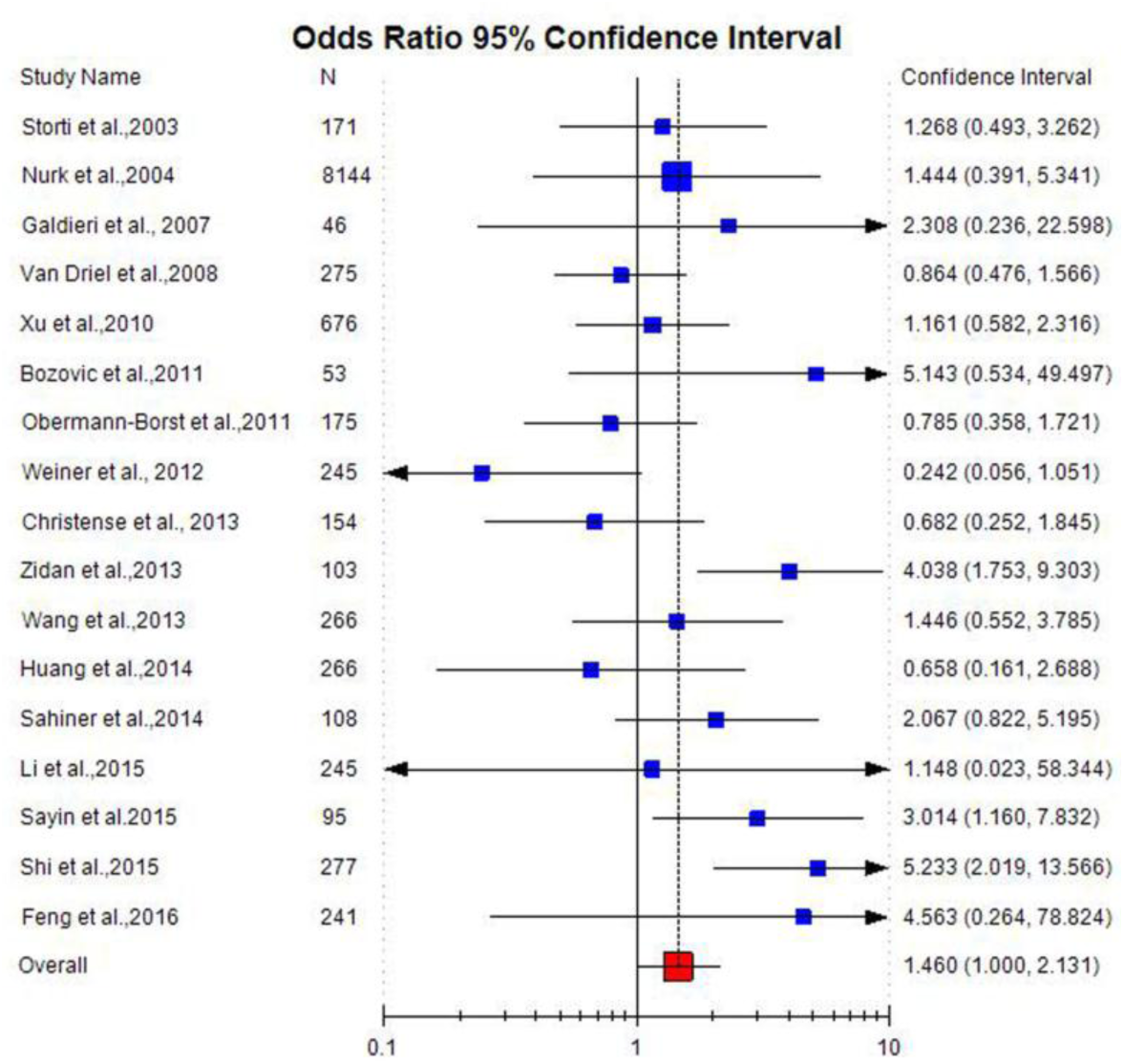
: Forest plots (random effect) of homozygote model (CC versus AA) showed significant association between maternal MTHFR A1298C polymorphism and CHD risk.

**Figure 3:**
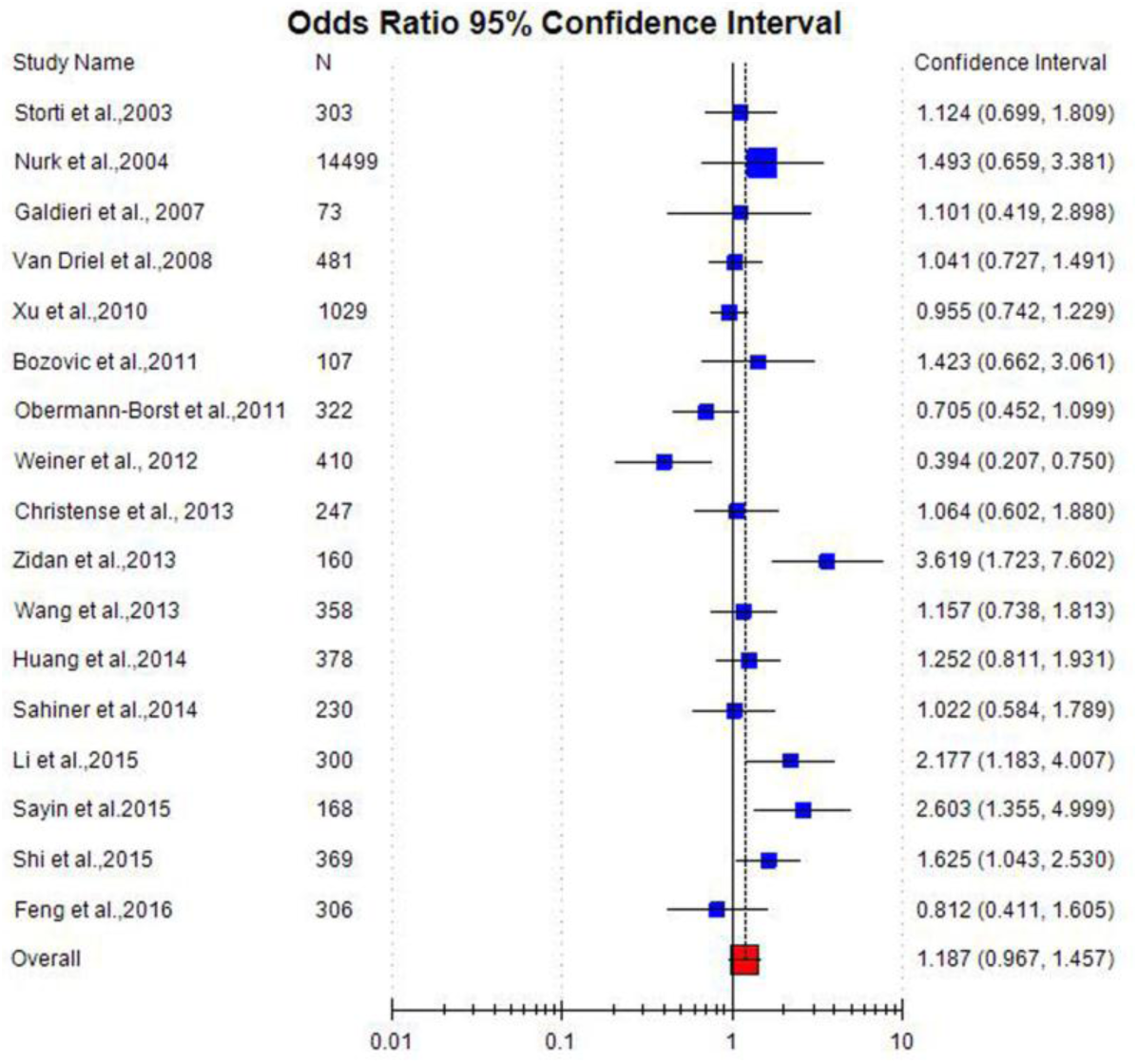
: Forest plots (random effect) of dominant model (CC + AC versus AA) showed significant association between maternal MTHFR A1298C polymorphism and CHD risk.

**Figure 4:**
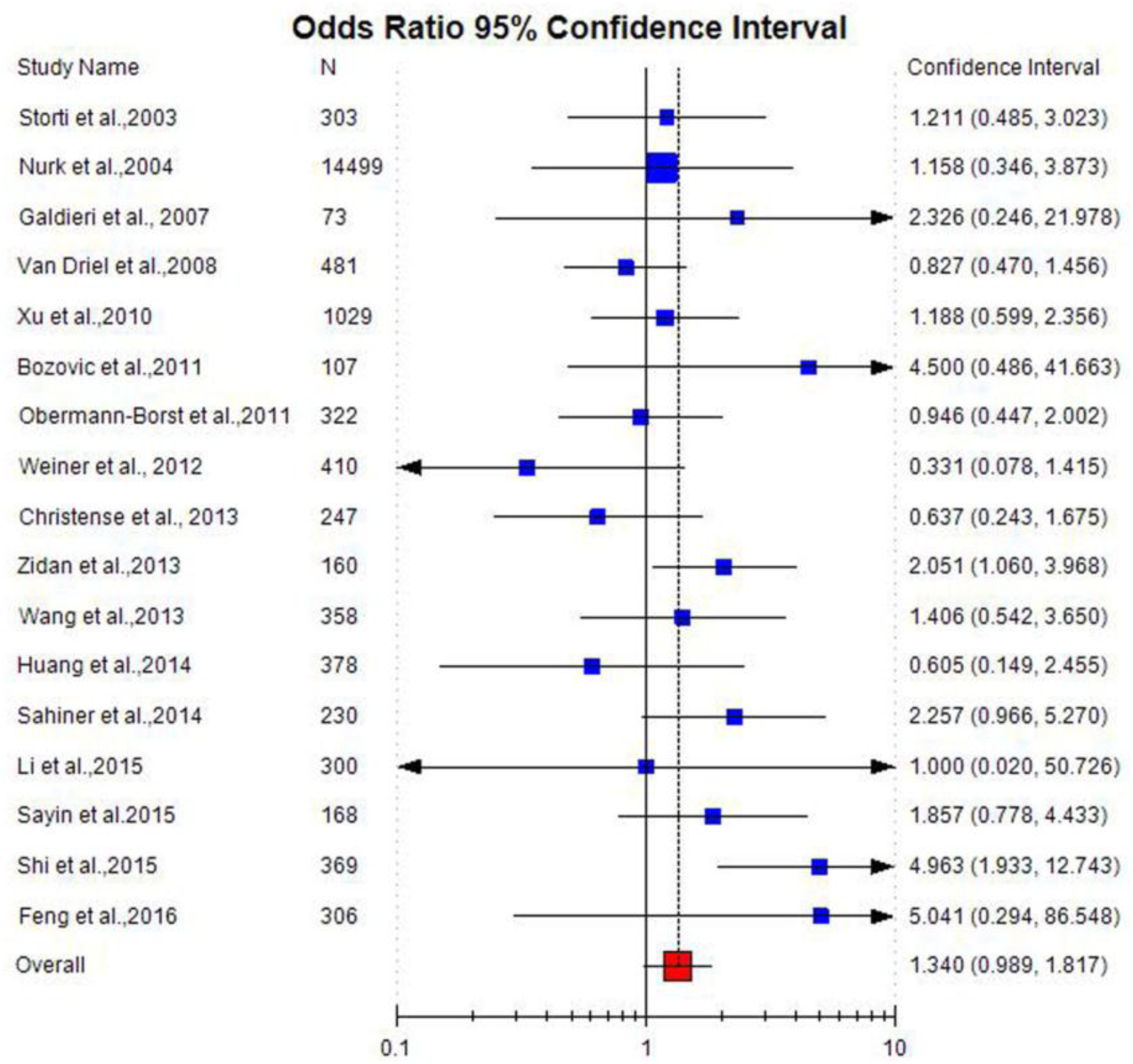
: Forest plots (random effect) of recessive model (CC versus AC+AA) showed significant association between maternal MTHFR A1298C polymorphism and CHD risk.

A insignificant low heterogeneity existed in between studies for recessive model (P_heterogeneity_ =0.06, I^2^= 37.8%), significant heterogeneity existed between studies for allele contrast (P_heterogeneity_ =0.0001, I^2^= 65.04%), dominant model (P_heterogeneity_ =0.0005, I^2^= 61.34%), high heterogeneity existed between studies for homozygote (P_heterogeneity_=0.005, I^2^= 53.68%,), and co-dominant (P_heterogeneity_ =0.006, I^2^= 52.23%,) comparisons.

### Publication bias

Funnel plots using standard error and precision values for allele and genotypes using random effect model were generated (Table 2; Figure 5,6). Symmetrical distribution of studies in the funnel plots suggests absence of publication bias except co-dominant model. This is also supported by Eggers test (p= 0.29 for C vs. A; p= 0.21 for CC vs AA; p= 0.21 for CC+AC vs. AA; p= 0.57 for CC vs. AC+AA and p= 0.04 for AC vs. AA) (Table 2).

**Figure 5:**
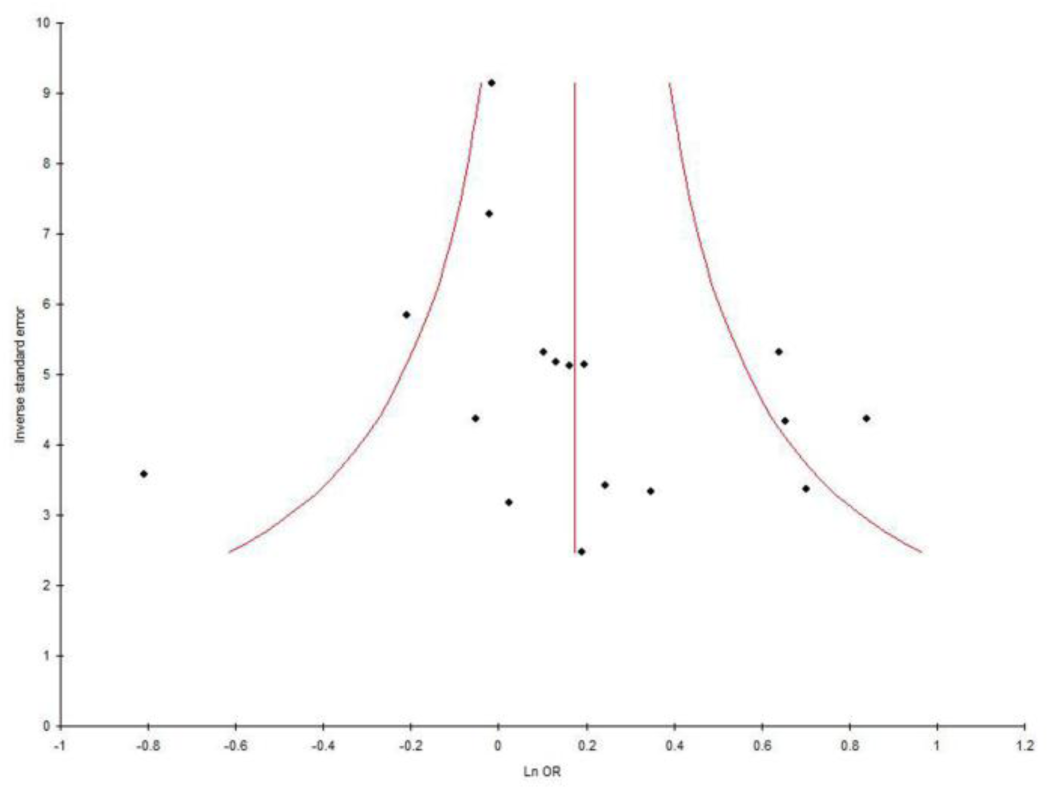
Funnel plots of precision by OR of MTHFR A1298C allele contrast.

**Figure 6.**
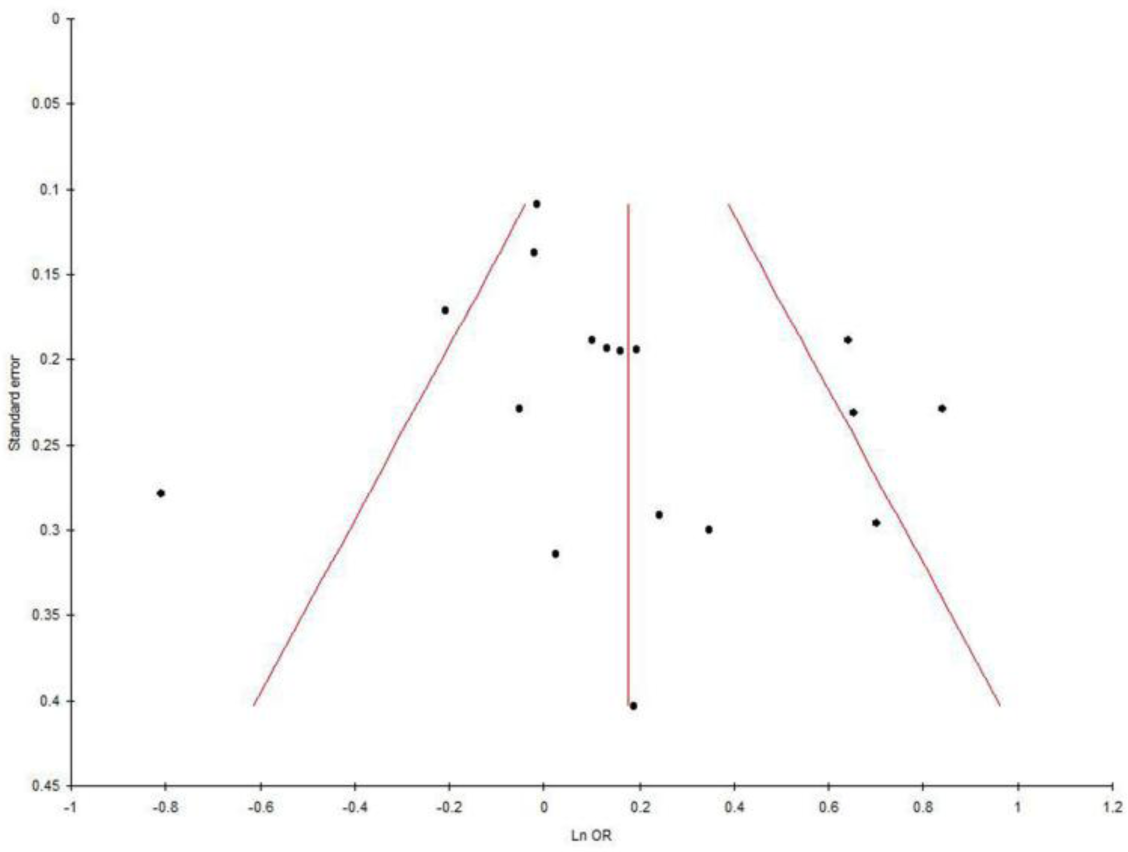
Funnel plots of standard error by OR of MTHFR A1298C allele contrast.

## Discussion

Normal MTHFR activity is crucial to maintain the pool of circulating folate and methionine and to prevent the accumulation of homocysteine (Frosst et al., 1995). Numerous case control studies have indicated an effect of the maternal MTHFR genotype rather than that of the affected child (Martinelli et al, 2001; Prescott et al, 2002) on fetal development. The association of MTHFR polymorphisms with the increased risk of CHD supports the protective effect of maternal use of folic acid with respect to the occurrence of CHD.

Folate deficiency and hypofunction of MTHFR enzyme activity were affected cell proliferation and apoptosis in developing fetus. Embryonic heart development is a complex process and proliferation and apoptosis play important roles (Fisher et al., 2000; Gittenberger-de Groot et al.,2005; Li et al., 2005). Inadequate proliferation or excess apoptosis can directly or indirectly result in CHD(Kruman et al.,2005). However, it is not clear whether folate deficiency alters proliferation/ apoptosis in the heart or not (Li et al.,2005). MTHFR gene knock out mice have reduced concentration of S-adenosylmethionine and increased concentration of S-adenosylhomocysteine, showed hyperhomocysteinemia and global DNA hypomethylation (Chen et al., 2001)

Meta-analysis is a statistical tool which is successfully used for combining results of several small, low powered case control studies to conclude the results with less statistical error and high statistical power. Numerous meta-analyses were published to confirm the small effect of gene polymorphism as risk factor for several disease like- cleft lip and palate (Rai,2014a,2018b); NTD (Yadav et al., 2015); male infertility (Rai and Kumar,2017d), Down syndrome (Rai,2011a; Rai et al.,2017c; Rai and Kumar,2018a); recurrent pregnancy loss (Rai,2016a), glucose 6- phosphate dehydrogenase deficiency (Kumar et al.,2016), bipolar disorder (Rai,2011b), Schizophrenia(Yadav et al.2016a, Rai et al.,2017e), autism (Rai,2016c; Rai and Kumar,2018c), Depression (Rai,2014c); Alzheimers disease (Rai,2016b,2017a); epilepsy (Rai and Kumar, 2018d), Uterine Leiomyoma (Kumar and Rai, 2018a), hyperurecemia (Rai,2016d), breast cancer (Rai,2014b; Rai et al.,2017b), esophageal cancer (Kumar and Rai, 2018b), lung cancer (Rai,2014d), prostate cancer (Yadav et al.,2016b),colorectal cancer (Rai,2015), digestive tract cancer (Yadav et al.,2018), endometrial cancer (Kumar and Rai, 2018c) and ovary cancer (Rai,2016e).

There are several limitations in current meta-analysis, which should be acknowledged- (i) unadjusted OR was used as association measure, (ii) only one gene polymorphism (MTHFR A1298C) was considered in present meta-analysis, (iii) only four databases were searched for studies, and (iv) gene- gene and gene -environmental interactions were not considered.

In conclusion, results of meta-analysis showed that maternal MTHFR A1298C polymorphism is a risk factor for congenital heart defects in fetus. Periconceptional intake of folate reduces the risk of congenital defects, because it donates methyl group for conversion of homocysteine to methionine and protect fetus from the teratogenic effect of higher concentration of homocysteine. Simultaneously methionine donates methyl group to S-adenosylmethionine for cellular methylation reactions in fetus and due to reduced MTHFR enzyme activity genomic hypomethylation occurs which affects gene expression in developing fetus. In future, larger studies from different regional as well as different ethnic population are required to find out the exact effect of maternal MTHFR A1298C polymorphism on fetal heart.

## Data Availability

All data are included in the manuscript

